# An atlas and evidence-based appraisal framework of psychiatric brain stimulation targets generated by causal network mapping

**DOI:** 10.1101/2025.02.25.25322842

**Authors:** Ryan D. Webler, Andrew R. Pines, Nicole Chiulli, Goncalo Cotovio, Ryan R. Darby, Jing Jiang, Juho Joutsa, Ningfei Li, Albino J. Oliveira-Maia, Shaoling Peng, Joseph J. Taylor, Shan H. Siddiqi

## Abstract

Causal network mapping is an emerging technique that can be used to derive optimal disorder/symptom-specific brain stimulation targets. This technique exploits incidental variability in brain lesion and brain stimulation locations, which creates a natural experiment in which causal inferences can be drawn between lesions or modulation of specific circuits and specific clinical outcomes. Circuits identified by causal network mapping, referred to as symptom-networks, represent candidate disorder/symptom-specific brain stimulation targets. The number of psychiatric symptom-networks has grown rapidly in recent years, creating a need for a comprehensive synthesis. To address this gap, this Resource presents an atlas of 12 psychiatric symptom-network targets and appraises them using an established evaluative framework. We describe how these targets can be localized with and without neuroimaging and highlight key considerations surrounding their trialing and implementation. These materials are designed to spur the translation of symptom-network targets and scaffold advancements in this quickly developing field.

## Introduction

Transcranial magnetic stimulation (TMS) has been investigated as a therapeutic intervention for psychiatric disorders since the 1990s and is currently the most commonly used brain stimulation technique in clinical practice (1–4). While its efficacy in depression is now well-established (5), establishing efficacy for other indications has been slow (6). This may be partly because the field has lacked an efficient process to generate and evaluate brain targets for new indications. Indeed, the main depression target has commonly been repurposed for other indications, often with disappointing results (7).

Recent landmark studies indicate that different psychiatric symptom clusters localize to distinct circuits, which may be useful for stratifying patients between existing targets (8,9) and to generate new ones (10). One class of methods specifically designed to generate new disorder/symptom-specific targets is causal network mapping. Causal network mapping uses normative brain circuit maps, usually generated using resting-state functional magnetic resonance imaging (rs-fMRI), to trace circuits connecting brain lesions or stimulation sites that selectively modify specific disorders/symptoms (11). Causal inference is possible due to a natural experiment in which lesion or stimulation sites incidentally vary in their location. Brain lesions incidentally emerge in different locations depending on the characteristics of a stroke, head injury, or other form of focal brain damage. Brain stimulation sites also incidentally vary in clinical practice due to variability or imprecision in the techniques used to apply stimulation; for instance, most TMS clinics use scalp measurements to approximate the stimulation target. When lesions or stimulation to a specific circuit induce a specific outcome, it is possible to infer a causal relationship between the circuit and the outcome. Lesions are more commonly used than stimulation sites due to the availability of large-scale data on patients who developed various symptoms after a stroke or other types of focal brain damage.

In principle, if damage to a particular brain circuit causes a symptom, then it may be reasonable to hypothesize that stimulating the same circuit would relieve that symptom. Data consistent with this possibility have emerged for different disorders, including depression (12), anxiety (13), tremor (14), both motor and cognitive symptoms in Parkinson’s disease (15,16), epilepsy (17), and vertigo (18). Across these disorders, lesions that incidentally cause a symptom map to a specific network, and TMS and/or deep brain stimulation (DBS) sites that incidentally target that network tend to modify that symptom. Broadly, this suggests that TMS and DBS may be prospectively applied to lesion-derived targets to reduce specific symptoms. Symptom-networks have been shown to outperform existing TMS target optimization strategies. For instance, TMS site connectivity to a depression symptom-network predicted depression reduction better than connectivity to the subgenual cingulate (19), the most common connectivity-based TMS target for depression (20).

Symptom-network targets have been generated at a much faster rate than they have been trialed (21–23) or implemented (24,25). A key factor impeding translation is the lack of a framework to synthesize and appraise this library of targets as it grows (26). The current Resource presents an atlas of 12 psychiatric symptom-networks, proposed TMS targets within each network, and guidance for how to target them with and without neuroimaging. We appraise the evidence supporting each symptom-network target using the GRADE system (Grading of Recommendations Assessment, Development, and Evaluation), a framework used by the World Health Organization, American College of Physicians, Cochrane Collaboration, UpToDate, and other major public health organizations to help clinicians make evidence-based decisions (27). We discuss how symptom-network targets can be translated and how the framework used to evaluate them can shape advancements in the rapidly developing causal network mapping field.

## Results

### Symptom-Network Atlas

Systematic search results are depicted in Supplementary Figure 1. The present atlas includes networks for 12 disorders/symptoms: addiction (28), aggression (29), anxiety (13), criminality (30), depression (19), emotional regulation (31), mania (32), obsessive-compulsive disorder (OCD)(33), pain (21), posttraumatic stress disorder (PTSD) (34), psychosis (35), and a transdiagnostic construct (36). Nine symptom networks were generated via lesion network mapping, one via DBS network mapping (OCD), and two via a combination of lesion, DBS, and TMS network mapping (depression and anxiety networks). Symptom-networks were standardized by correlating them with a normative functional connectome (37). For each voxel in the brain, we computed a spatial correlation to quantify the similarity between that voxel’s connectivity profile and the network in question (38). This analysis yields a map depicting the extent to which each voxel, if stimulated, would be expected to target the corresponding network (Figure 1). Results were normalized (i.e., Z-scored) to facilitate between-network comparison.

**Fig. 1.**
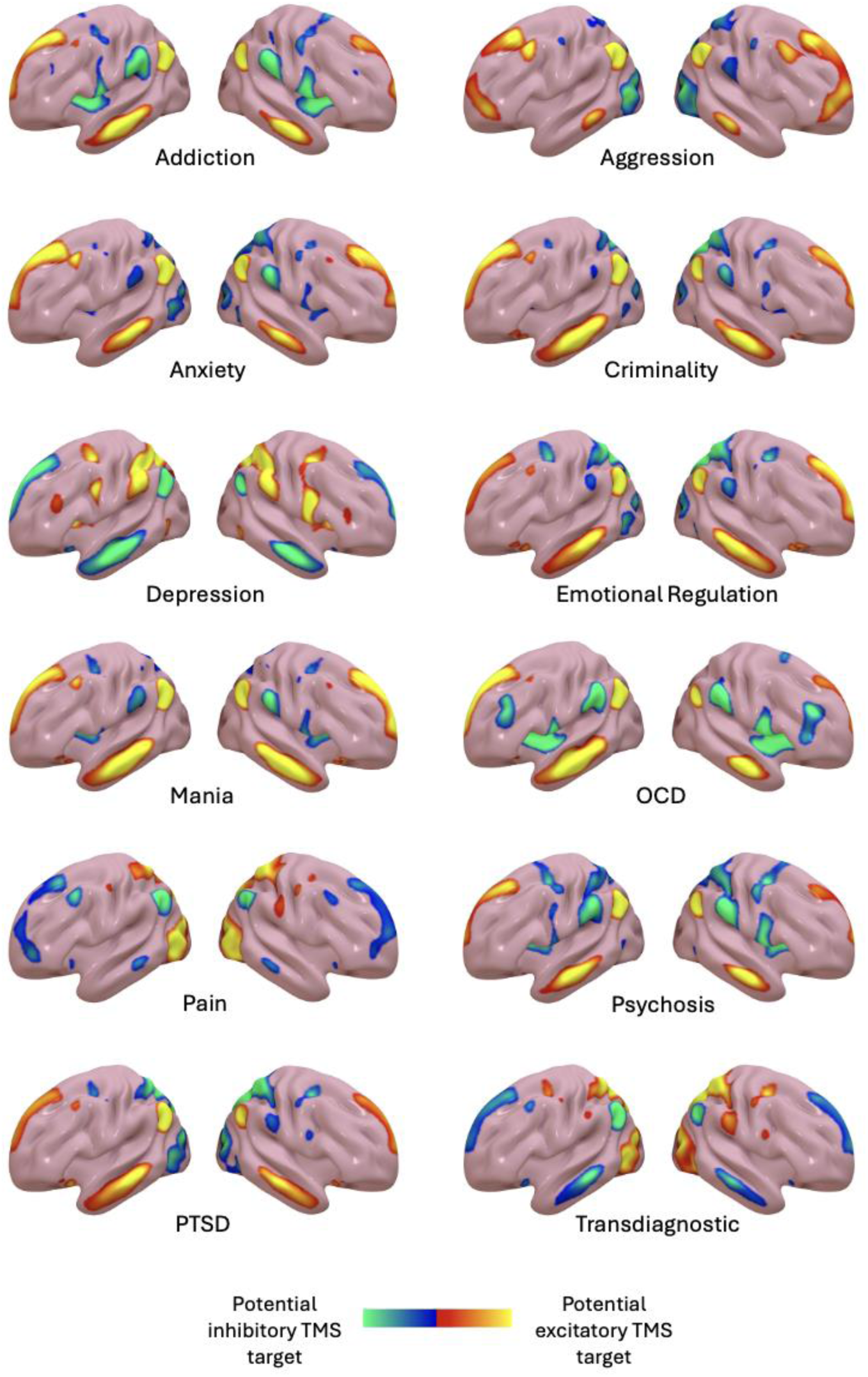
Symptom-network atlas. Whole-brain maps depicting the voxels whose connectivity profile best resembles each respective symptom-network. Depicted networks were thresholded at |Z| > 1.5. Positive connections of all networks represent potential excitatory TMS targets; negative connections represent potential inhibitory TMS targets.

Lesions and DBS presumably disrupt activity in affected brain areas and their positive connections. If disrupting network activity causes a symptom, then facilitating network activity should reduce it; thus, it is plausible that positive connections of lesions/DBS sites that cause a symptom may represent potential excitatory TMS targets (12,13). In contrast, negative connections of lesions/DBS sites that cause a disorder/symptom or positive connections of lesions/DBS sites that protect against or reduce a symptom may represent potential inhibitory TMS targets. However, it is important to note that the optimal stimulation paradigm is an empirical question that requires testing. Consistent with the above facilitation/disruption hypotheses, a recent study showed that lesions that cause depression and brain stimulation sites that relieve it map to a common network (19). As predicted, putatively facilitative excitatory TMS and disruptive DBS stimulation sites yielded inverted results. Similar results have also been seen for anxiety (13). In Figure 1, to facilitate visual comparison across sign inversions, networks generated by lesions/DBS sites that protect against or reduce a symptom were inverted in the maps below.

### Relative Symptom-Network Signatures

Several symptom-networks depicted in Figure 1 appear to share similar cortical signatures. To compare symptom-networks, we performed a winner-take-all analysis which parcellated the brain into regions most associated with each individual symptom network. Each voxel in the prefrontal cortex (PFC) was assigned to a symptom-network with which it showed greatest connectivity (Figure 2). We restricted our focus to positive PFC connections given that the vast majority of clinical TMS studies have applied excitatory TMS to PFC target spaces, so safety is well-established. Moreover, excitatory TMS incidentally applied to positive connections of lesions that cause a disorders/symptom has been shown to reduce that same disorder/symptom (12,13). The effect of inhibitory TMS on negative connections of disorder/symptom causing lesions remains uninvestigated.

**Fig. 2.**
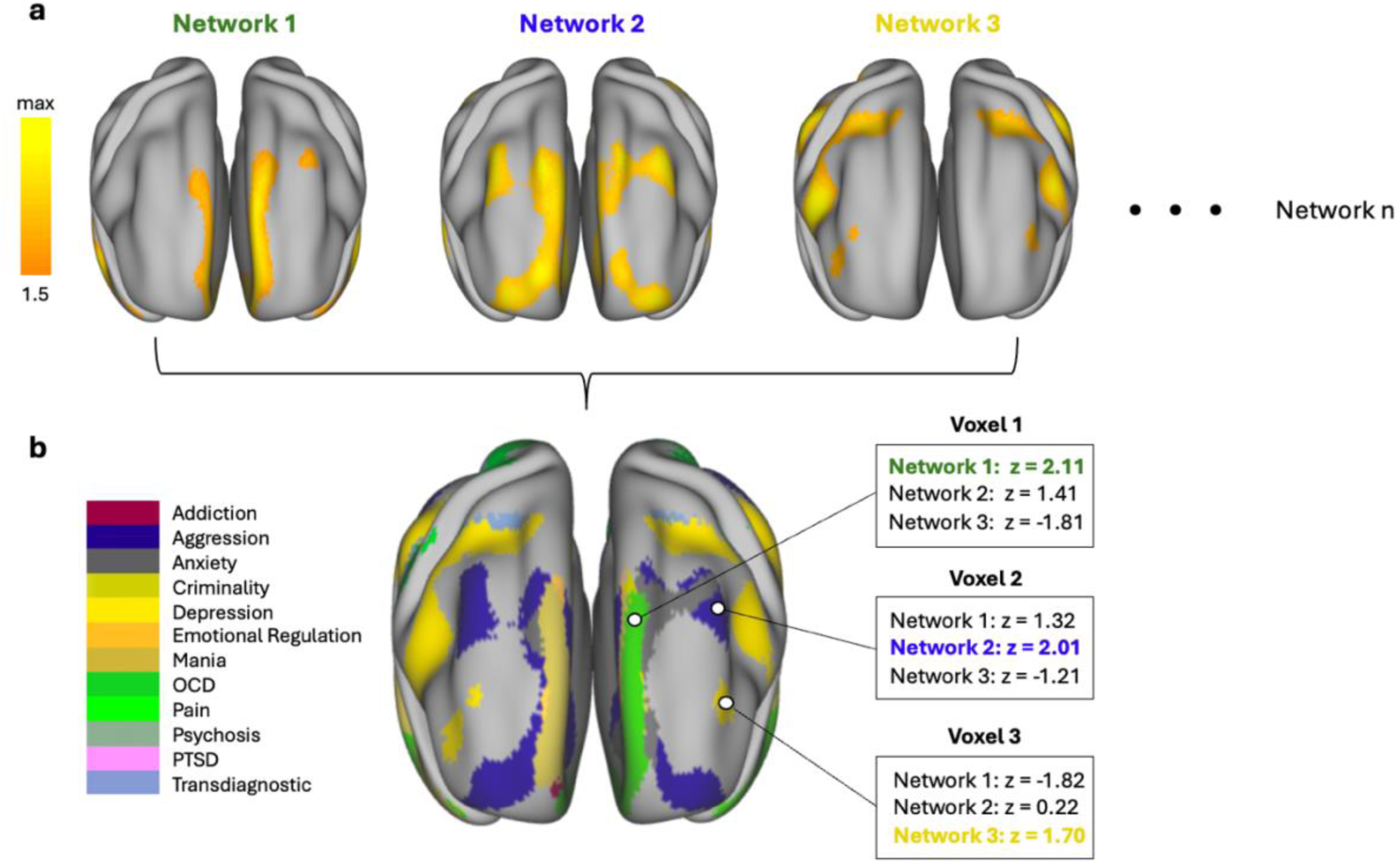
Winner-take-all map. Thresholded symptom-networks (a) combined into a winner-take-all map in which each voxel was labeled with the most strongly associated network; (b) z-values for three different networks at three different voxels are shown. Networks 1, 2, and 3 had the highest z-value at voxels 1, 2, and 3, respectively. Therefore, voxels 1, 2, and 3 were associated with networks 1, 2, and 3, respectively.

The winner-take-all map generated from this analysis demonstrates that different symptom-networks have unique relative positive connectivity signatures in the medial, lateral, and orbitofrontal PFC. Consistent with findings that excitatory left dlPFC TMS may be more effective for depression than other indications, the depression network showed the strongest positive connectivity to the left dlPFC.

### Proposed Symptom-Network TMS Targets

The relative network signatures depicted in Figure 2 reveal PFC locations most strongly positively connected to each network *compared* to other networks. However, other PFC locations may be more strongly connected to a given symptom network – even if they are relatively more connected to another network. Stimulating locations that are most connected to a given network should have the greatest impact on the overall network. Therefore, our proposed symptom-network targets are mostly localized to PFC connectivity peaks.

Proposed symptom-network targets are depicted alongside common locations used for TMS targeting in clinical practice in Figure 3 and their MNI coordinates are listed in Table 1. Scalp coordinates from newly developed, high-precision coordinate systems (continuous proportional coordinate space, CPC; Tetra codes) are also provided to facilitate implementation for the vast majority of cases in which neuroimaging-guided targeting is unavailable. CPC and Tetra codes transform 3-dimensional MNI brain coordinates into 2-dimensional scalp-atlas sites that can be manually marked or printed on commonly used scalp caps to guide TMS targeting. Details on how to implement these approaches can be found in two recent open-access publications (39,40).

**Fig. 3.**
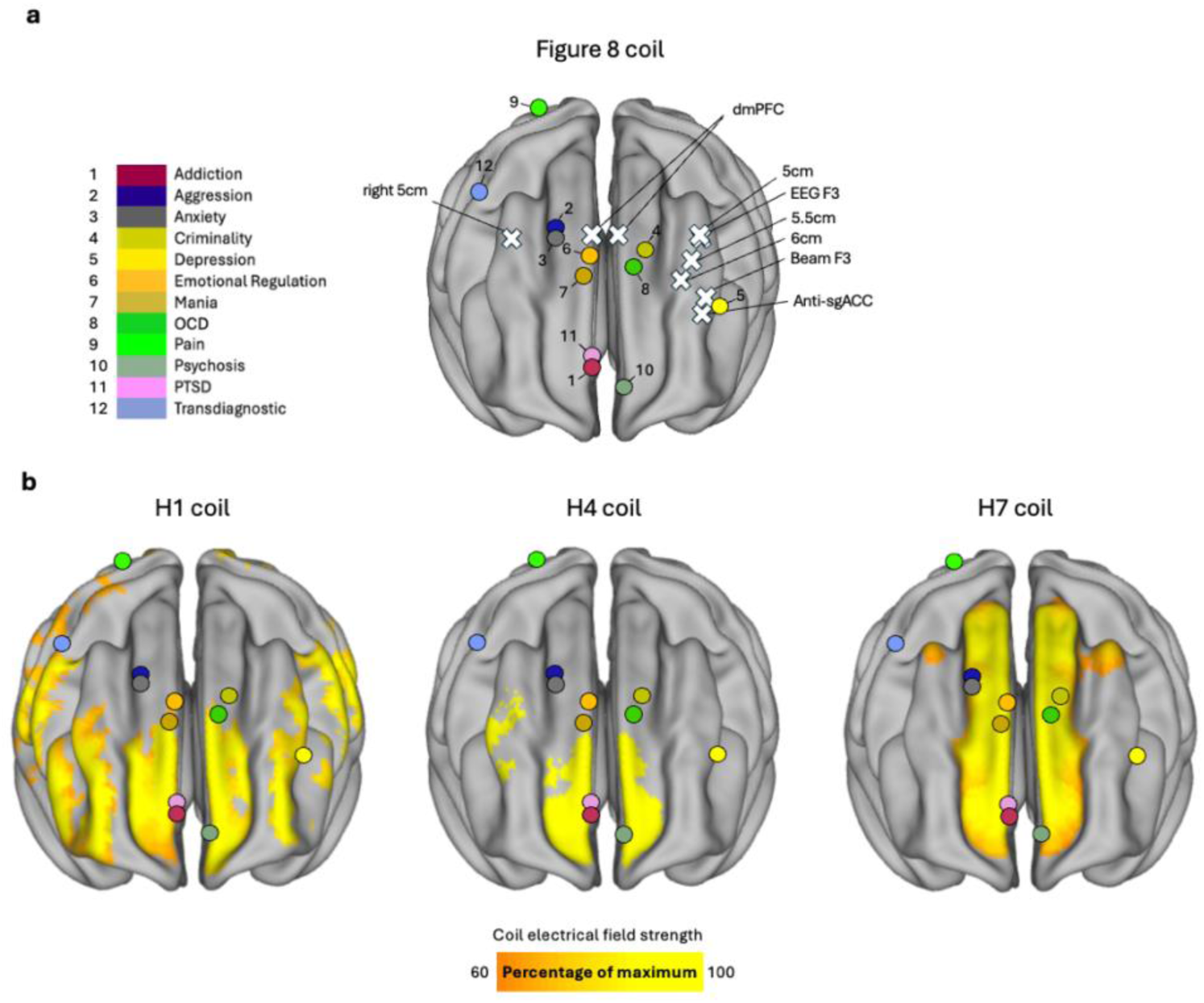
Symptom-network targets. a. Proposed symptom-network targets labeled in color coded circles and several common locations for Figure-8 coils. b. Proposed symptom-network targets overlaid on electric-fields of H-coils used to treat depression (H1 coil), addiction (H4 coil), and OCD (H7 coil). The H1 coil e-field was calculated using SimNIBS, while the H4 and H7 coil e-fields were extracted from a prior publication (28). E-fields were standardized such that the max voxel was set to 100 and all other voxels set to a percentage of the max voxel.

As depicted in Figure 3, most proposed symptom-network targets are relatively distant from common Figure-8 coil targets used to treat depression. Several are also outside the e-field of multiple H-coils used to treat depression, OCD, and addiction, indicating that re-orientation is likely required to stimulate these targets even when using broadly targeting coils.

**Fig. 4.**
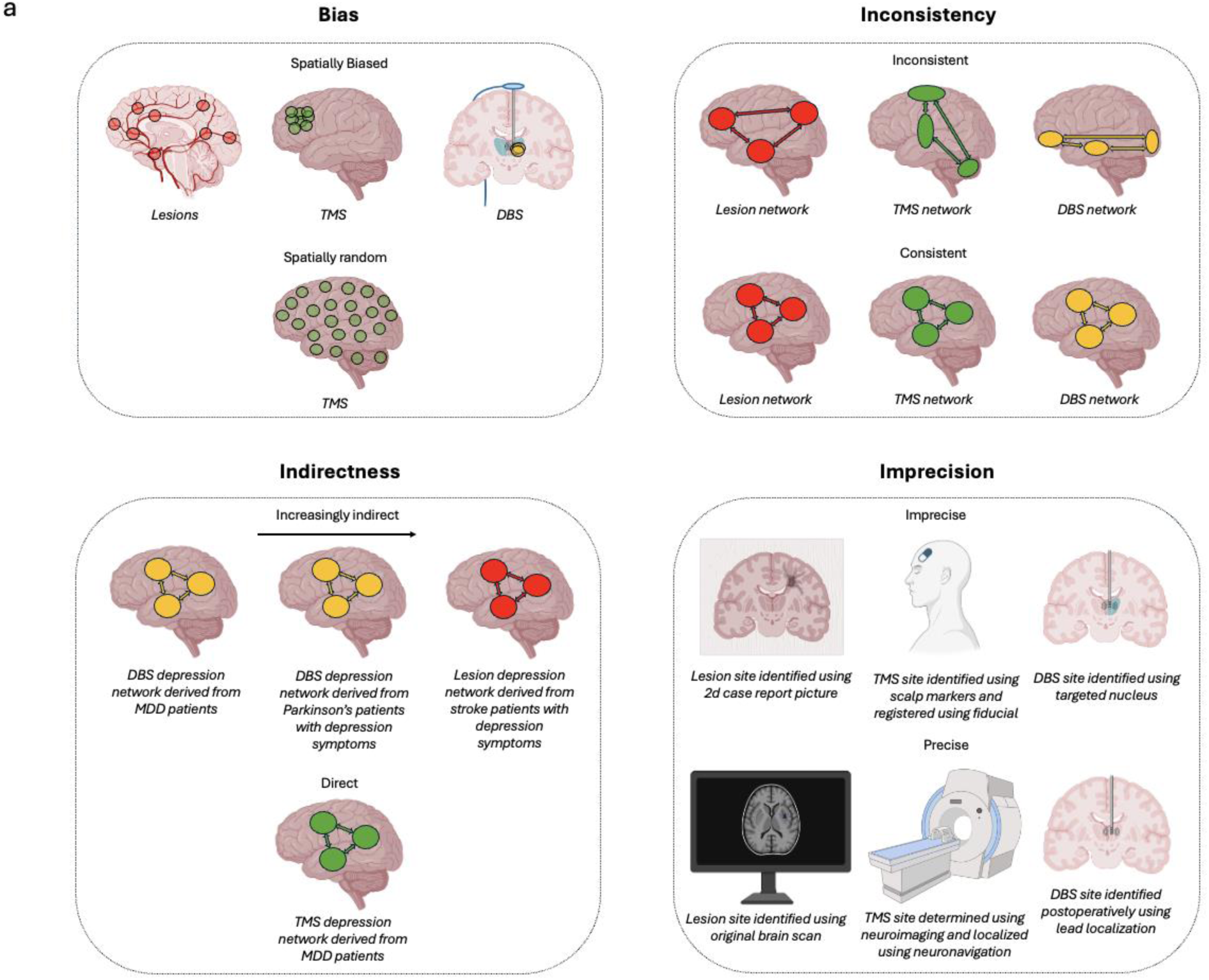
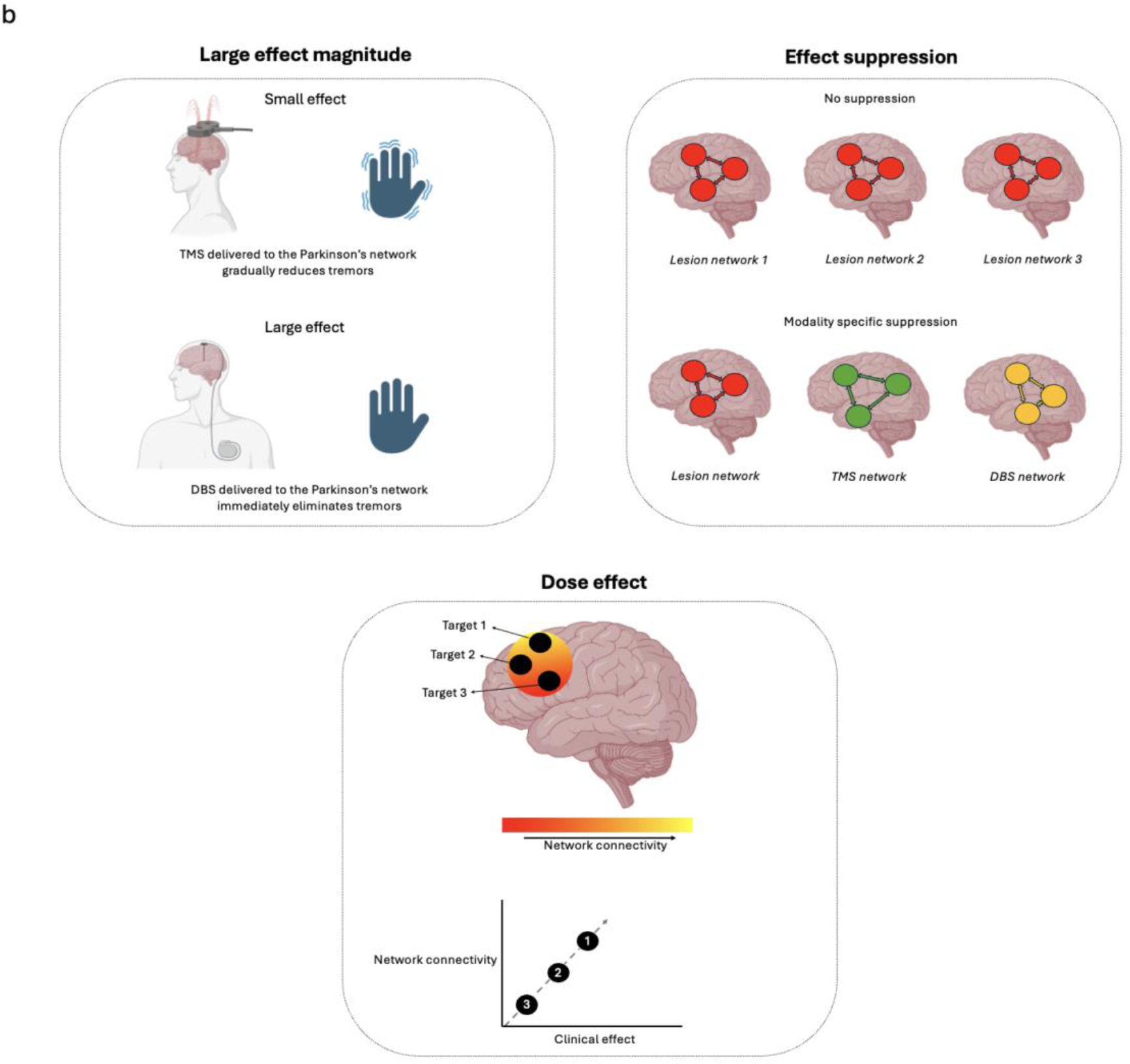
GRADE criteria. *a. Evidence weakening criteria.* <u>Bias:</u> Incidentally varying lesions, TMS, and DBS sites are pseudorandomly distributed, introducing spatial bias. Lesions are associated with the least spatial bias, as they are more likely to occur near blood vessels but are spatially diffuse. TMS is associated with more spatial bias, as it is applied to a limited number of target spaces. DBS is associated with the most spatial bias, as it is applied within small sub-nuclei. Prospectively targeting random locations with TMS would eliminate spatial bias. <u>Inconsistency:</u> Inconsistency is present if a disorder/symptom maps to different networks using different causal modalities. Consistency can be established by convergence across causal modalities. <u>Indirectness:</u> A symptom-network derived for the purpose of generating an optimal TMS target for MDD would be slightly indirect if generated from DBS sites in MDD patients, more indirect if generated from DBS sites in Parkinson’s patients with depression symptoms, and most indirect if generated from lesion sites in stroke patients with depression symptoms. A network generated from TMS sites in MDD patients would be most direct. <u>Imprecision:</u> Lesions localized using 2d case report images, TMS sites identified using scalp markers and retrospectively localized using a fiducial marker, and DBS sites localized to a sub-nucleus are imprecise. Lesions localized using original brain scans, TMS sites determined using neuroimaging and localized using neuronavigation, and DBS sites localized postoperatively using lead localization are precise. *b. Evidence strengthening criteria.* <u>Large effect magnitude:</u> TMS delivered to the Parkinson’s network may generate a small effect that results in the gradual reduction of tremor symptoms. DBS delivered to the Parkinson’s network may generate a large effect that results in the immediate elimination of tremor symptoms. <u>Effect suppression:</u> Different causal methods introduce unique sources of error which may suppress effects, as depicted by small differences in networks generated by lesions, TMS, and DBS. <u>Dose effect:</u> A significant relationship between brain stimulation site connectivity and clinical effects indicates a dose effect. As depicted here, stronger TMS site connectivity to the network (connectivity increases from red, to orange, to yellow) yields greater clinical effects.

**Table 1.** Symptom-network target coordinates. Optimal symptom-network target MNI, continuous proportional coordinate space (CPC), and Tetra code coordinates are depicted.

| Indication | MNI |  |  | CPC |  | Tetra |
| --- | --- | --- | --- | --- | --- | --- |
|  | X | Y | Z | Pnz | Pal | Code |
| Addiction | 0 | 68 | 8 | 0.144 | 0.497 | 49FW |
| Aggression | 26 | 30 | 54 | 0.353 | 0.594 | 86HV |
| Anxiety | 23 | 34 | 52 | 0.336 | 0.584 | 7XHM |
| Criminality | -14 | 40 | 50 | 0.317 | 0.443 | 7HCP |
| Depression | -43 | 37 | 24 | 0.276 | 0.316 | 6P82 |
| Emotional Regulation | 13 | 43 | 49 | 0.306 | 0.547 | 79H2 |
| Mania | 16 | 39 | 51 | 0.321 | 0.559 | 7MH6 |
| OCD | -11 | 45 | 47 | 0.297 | 0.454 | 76CX |
| Pain | 27 | -55 | 61 | 0.666 | 0.599 | HMJ3 |
| Psychosis | 0 | 66 | 8 | 0.144 | 0.497 | 49FW |
| PTSD | 1 | 64 | 12 | 0.161 | 0.499 | 4MFW |
| Transdiagnostic | 47 | -7 | 53 | 0.469 | 0.674 | C6MH |

### Symptom-Network TMS Target Grades

Symptom-network target evaluations are shown in Figure 5. Here, we use the depression network target as an example of how these evaluations were performed. Explanations for other symptom-network evaluations can be found in Supplementary Materials.

**Fig. 5.**
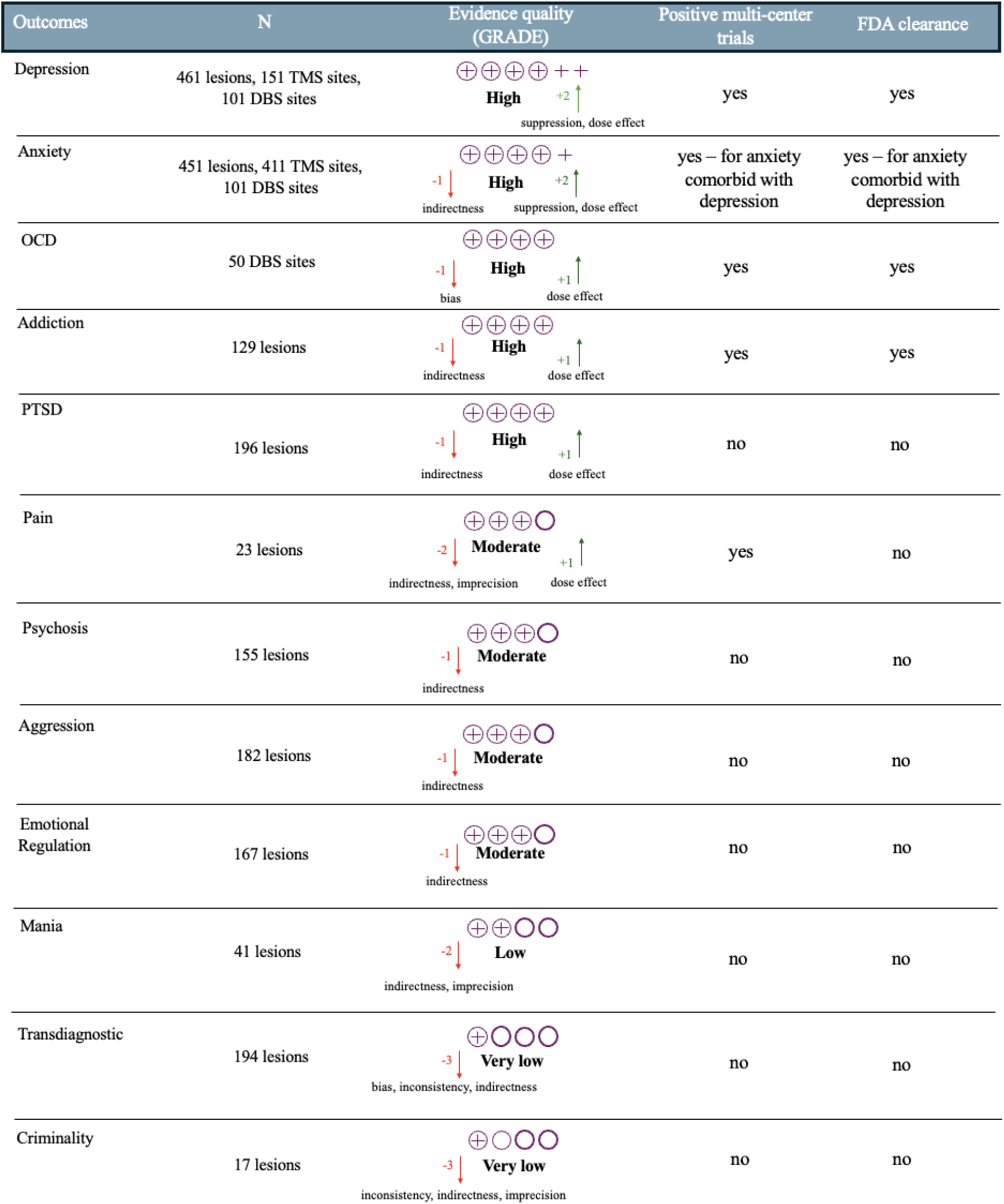
Symptom-network target grades. Each symptom-network target was evaluated on the basis of the GRADE criteria and given a composite score. Consistent with the GRADE framework, these grades are defined as the following. <u>High quality</u>: Further research is very unlikely to change our confidence in the estimate of effect. <u>Moderate quality</u>: Further research is likely to have an important impact on our confidence in the estimate of effect and may change the estimate. <u>Low quality</u>: Further research is very likely to have an important impact on our confidence in the estimate and is likely to change the estimate. <u>Very low quality</u>: We are very uncertain about the estimate.

Of the network targets described here, the depression network target performs best against the GRADE criteria. <u>Bias:</u> The depression network target was not downgraded for bias, as the effects of key confounders including lesion size and overall symptom severity were accounted for. Moreover, spatial bias was limited by the inclusion of spatially heterogeneous lesion sites and brain stimulation sites spanning multiple targets. <u>Inconsistency:</u> It was not downgraded for inconsistency due to significant spatial correlations between the lesion, DBS, and TMS networks that comprise it. <u>Indirectness:</u> Although multiple data types and diagnoses were included in the depression network, it was not downgraded for indirectness because it included data from over 150 patients receiving TMS for MDD. <u>Imprecision:</u> It was not downgraded for imprecision, as it was derived from several large sample datasets that generally used high resolution techniques to localize lesions/brain stimulation sites. <u>Large effect magnitude:</u> The clinical effect sizes were relatively small, so the network was not upgraded for effect size. <u>Effect suppression</u>: It was upgraded for suppression, as the measured effect size was likely an underestimate due to integration of multiple causal data-sources with unique confounders. <u>Dose effect:</u> It was upgraded for dose-effect, as greater connectivity to the depression network predicted greater clinical improvement with therapeutic brain stimulation. <u>Overall grade:</u> The evidence supporting the depression network was evaluated as high quality.

## Discussion

The current Resource includes 12 psychiatric symptom-networks, proposed TMS targets for each network, and a GRADE-informed evaluation of these targets. These materials can help clinicians locate symptom-network targets for approved indications and decide which of these targets to stimulate for a given patient on the basis of clinical need and the current evidence-base. They can also inform clinical trials and guide advancements in causal network mapping methods.

In the United States, TMS is an FDA-cleared treatment for depression, OCD, addiction, and anxiety comorbid with depression. Proposed symptom-network targets for these indications fall within FDA-cleared target spaces and were evaluated as high quality. Therefore, they may be considered for direct clinical implementation. In clinics where neuroimaging is available, a brief structural MRI (i.e., 6-minute T1) can be collected, target coordinates can be transformed to individual patient space, and neuronavigation can be used to orient to these coordinates in each patient. In cases where advanced neuroimaging (e.g. resting-state functional MRI) is available, the targets may be further personalized based on individualized connectivity (34). When neuroimaging is unavailable, scalp landmarks can be used to approximate target locations (41).

While this comes with a tradeoff in precision, it enables more widespread implementation, as most TMS clinics currently identify targets using scalp measurements. For example, the depression target can be approximated using the Beam F3 method, which is commonly used in clinical practice and has been employed by many clinical trials (42,43). Newly developed scalp coordinate systems such as CPC and Tetra codes can also be used to target symptom-networks (see Table 1), which may increase precision compared to standard scalp targeting approaches. Adoption of these systems is supported by open-access resources (39,40).

Diffuse H-coils (i.e., H1, H4, and H7) also overlap with some proposed targets. While these coils are less precise, they stimulate a larger brain volume, which may be useful for network-level stimulation. H-coils also carry the added benefit of being supported by multi-center clinical trials with associated FDA clearances for specific indications. The H1 coil, which overlaps with the depression, addiction, OCD, PTSD, psychosis, and mania targets, is cleared by the FDA for MDD. The H4 coil, which overlaps with the addiction, PTSD, and psychosis targets, is cleared for nicotine use disorder. The H7 coil, which overlaps with the OCD target, the addiction target, and several others, is cleared for OCD. Each of these clearances was supported by FDA-registered multi-center clinical trials. The H7 coil is also supported by a multi-center clinical trial for alcohol use disorder (44), but is not cleared by the FDA for this indication at the time of this writing.

In complex cases where multiple FDA-cleared TMS indications are present, clinicians could balance considerations of evidence strength and clinical need (i.e., acute risk/distress) and work collaboratively with patients to determine which symptom-network target to stimulate. For example, a patient presenting with depression, OCD, and substance use disorder might be a candidate for at least three symptom-network targets. If all disorders/symptoms are equally severe, the depression target might be favored because it currently has the highest evidence quality. If the clinician believes that the patient’s OCD is most severe and/or is driving the other symptoms, then the OCD target might be recommended. If the patient feels that their substance use is causing the most acute distress, then the addiction target might be chosen due to the patient’s stated priorities.

Clinicians may be tempted to target multiple symptom-networks for FDA-cleared indications during the same treatment course. However, this adds an untested variable to the treatment, creating potential interaction effects between multiple targets. For instance, stimulating multiple networks may produce iatrogenic push-pull effects that counteract each other. This possibility is supported by at least one randomized trial in which motor cortex TMS significantly reduced motoric Parkinson’s symptoms relative to sham, but motor cortex TMS combined with dlPFC TMS did not (45). These and other findings showing that TMS can unintentionally worsen some symptoms (46) support the prioritization of single-target TMS protocols with established safety/efficacy over currently unsupported multi-target protocols.

However, multi-target protocols may still become valuable in the future with clinical trials that carefully account for added variables introduced by multi-target protocols, such as the correlation between the targeted networks, the time interval between targets to avoid disturbing immediate post-stimulation homeostatic effects, the optimal order of targets, the ideal stimulation parameters for each target, and other factors. Due to these open questions, at the current time we can only recommend stimulating one target at a time.

Clinical decision-making should be shaped by the accumulation of new evidence, which will likely lead to changes in current symptom-network target ratings. For example, the depression network target could be prospectively trialed against another network target not linked to depression. A positive result would further reduce bias and strongly demonstrate specificity. A negative result from a trial like this, or from future causal network mapping analyses, could lead to the downgrading of a given network target. New evidence will also likely lead to the inclusion of additional symptom-networks generated from lesions and brain stimulation methods including TMS and DBS, as well as by newer methods such as transcranial focused ultrasound (tFUS). The addition of network targets generated by new methods, and for use by those methods, will require independent appraisal using criteria like those presented here.

Clinical trials of current and future symptom-network targets are necessary. If safety and efficacy have not yet been established, sham-controlled trials are critical. For established indications, head-to-head clinical trials are still valuable to compare distinct targets for the purpose of target optimization. Current optimal symptom-network TMS targets are generally restricted to the PFC given that the vast majority of psychiatric clinical TMS trials have stimulated PFC target spaces and safety and efficacy/effectiveness data are therefore strongest for PFC targets. However, symptom-networks are distributed and include many brain areas outside the PFC. Future clinical symptom-network trials should compare proposed PFC network targets against non-PFC target locations. For example, a future comparative depression network trial could test the well supported dlPFC network location against the depression network peak located in the inferior parietal lobule.

Causal network mapping may yield superior brain stimulation targets compared to other approaches but current symptom-networks have noteworthy limitations that could be redressed by methodological advancements consistent with the standards outlined here. For example, variance in lesion and stimulation site location is incidental but not entirely random. Lesions occur throughout the brain but are more likely to occur in certain areas (e.g., blood vessels in the case of stroke); brain stimulation sites cluster around a small number of common target spaces. While specificity analyses help to control for this bias, the use of prospective experimental designs that apply TMS to random locations within established target spaces would mitigate it. Another limitation is the use of questionnaires (sometimes single questionnaire items) with questionable reliability/validity for measuring a specific construct that is encoded in the brain. Imprecision introduced by poor behavioral measurement could be addressed through the use of more reliable and valid psychometric batteries. Leveraging individualized rs-fMRI could further decrease imprecision by accounting for individual differences in functional connectivity (47) that are obscured by group-level connectomes. Another limitation is potential publication bias, which has not been formally assessed in this literature, and could be mitigated by pre-registration. Finally, it must be acknowledged that the vast majority of causal network mapping studies have been published by the group that includes authors of this Resource. The expansion of causal network mapping to new research groups and the application of methodological advancements consistent with the framework described here could significantly strengthen the field.

In conclusion, the present Resource is designed to support the translation of symptom-network targets which has been slowed by the lack of a consensus resource. The GRADE-informed evaluative framework presented herein is intended to provide a set of standards that guide clinicians to implement symptom-network targets when appropriate and researchers to trial current targets and generate new ones.

## Methods

### Symptom-Network Search Criteria

Psychiatric symptom networks were identified via a systematic search (performed by RW and AP) on PubMed, Web of Knowledge, and Embase that used the following terms: (‘lesion’ OR ‘brain lesion’ OR ‘deep brain stimulation’ OR ‘DBS’ OR ‘transcranial magnetic stimulation’ OR ‘TMS’) AND (‘circuit mapping’ OR ‘network mapping’ OR ‘lesion network mapping’ OR ‘causal network mapping’ OR ‘convergent causal mapping’ OR ‘common brain circuit’ OR ‘common brain network’ OR ‘network target’) AND (‘psychiatric’ OR ‘neuropsychiatric’ OR ‘psychiatric symptoms’ OR ‘neuropsychiatric symptoms’). When multiple networks existed for a given disorder/symptom (e.g., two depression networks), the network generated from the larger dataset was selected. To promote inclusivity, psychiatric disorders/symptoms were defined liberally to include any disorder/symptom linked to a Diagnostic and Statistical Manual of Mental Illness (DSM-5) classification. For example, criminality and aggression were included as they are correlates of antisocial personality disorder (48,49), a DSM-5 classification. Pain was included because it is a common psychiatric comorbidity (50) and a common manifestation of somatic symptom disorders (51).

### Symptom-Network Comparison

Networks were standardized by correlating them with a normative functional connectome that estimates the functional connectivity of each voxel to every other voxel in the brain. This database is comprised of functional connectivity data from 1,000 healthy young adults (37). For each voxel in the brain, we computed a spatial correlation to quantify the similarity between that voxel’s connectivity profile and the network in question (38). Results were Z-scored to facilitate between-network comparison. Relative network signatures were identified via a winner-take-all analysis that assigned each voxel to the symptom-network it is maximally connected to. We focus here on positive prefrontal cortex (PFC) connections because most clinical TMS studies have used excitatory TMS to stimulate PFC targets.

### Symptom-Network Targets

In addition to standardizing symptom networks, correlating each symptom-network with the normative connectome revealed locations with strong connectivity to each network which represent candidate TMS targets. Proposed targets were generally PFC voxels that showed the strongest positive connectivity to each symptom-network. To examine the effect of different TMS coils on these targets, we plotted them on a brain surface alongside several common Figure-8 depression targets as well as the estimated electrical-field (e-field) current of three deep TMS coils used to treat depression, OCD, and addiction.

### Evaluating Symptom-Networks

Two authors (RW and AP) independently evaluated symptom-networks using the GRADE framework. Consensus was then reached and final evaluations were reviewed and approved by the senior author (SS). GRADE includes eight factors that weaken or strengthen evidence quality (52). Factors that weaken evidence quality are (1) study bias, (2) inconsistency, (3) indirectness, (4) imprecision, and (5) publication bias. Factors that strengthen evidence quality are (6) large effect magnitude, (7) study design features that likely suppress effect magnitude, and (8) demonstrated dose-response gradient.

Study bias refers to methodological limitations that increase the probability of a misleading outcome. Bias is reduced when confounders are eliminated; the gold standard for unbiased studies is a randomized controlled trial, in which the intervention is the only factor that systematically differs between groups. This is a useful way to minimize bias when comparing a TMS target to sham, but not when seeking to identify a new target, as it is impractical to compare every possible TMS site in a randomized trial. Causal network mapping addresses this retrospectively by contrasting lesion and brain stimulation sites that cause or modify a symptom against those that do not (11). A strength of this approach is that it contrasts many lesion/brain stimulation sites simultaneously, increasing the probability that an optimal target will be identified. A weakness is that these sites are only pseudorandomly distributed, as lesions occur more frequently in certain brain areas (i.e., near blood vessels) and brain stimulation sites cluster within a small set of common targets. This inherently biases significant results to specific brain areas and their connections. Spatial bias is lower with lesions, which occur throughout the brain, more apparent with TMS, which is applied to individual target spaces, and most apparent with DBS, which is applied to specific subnuclei. Overall, spatial bias introduced by these methods is less than in typical brain stimulation RCTs which typically restrict their focus to a single target. We therefore did not generally downgrade symptom-networks for bias if they included control analyses that: (1) accounted for confounders such as lesion size and overall symptom severity, and (2) demonstrated that the network effects were specific to the given disorder/symptom. However, maps generated by DBS sites alone were downgraded due to the limited spatial variability of DBS sites and the spatial bias this causes.

Inconsistency refers to heterogeneity of an effect across studies/methods. Symptom-network consistency is typically demonstrated via spatial correlations between two halves of the same data-set (split-half consistency), different datasets using the same causal method (e.g., multiple lesion datasets), or different datasets using a different causal method (e.g., lesion and TMS datasets). A significant spatial correlation between symptom-networks generated using different causal methods is a particularly strong indicator of consistency given that confounders from one method may be mitigated by other methods. Spatial correlations between symptom-networks and high-quality neuroimaging results (i.e., meta-analytic or consortium level) are also commonly tested, even though these results are correlative rather than causal. We downgraded symptom-networks for inconsistency if significant within (i.e., split-half) or out-of-sample network similarities were not established.

Indirectness is the extent to which the intended use of a target differs from the published studies on that target. The most direct symptom-networks are those that are derived from the same symptom/diagnosis being treated and the same method being applied. For example, a depression map derived for the purposes of optimizing a TMS target for major depressive disorder (MDD) is most direct if generated from participants with MDD using TMS network mapping. A depression map generated for the same purpose using LNM, or from a transdiagnostic sample with depression symptoms, is less direct. We downgraded symptom-networks for indirectness if they were derived purely from lesion data or samples that were significantly different from the target population.

Imprecision refers to factors that widen the confidence interval around an effect. Larger sample sizes improve the precision of symptom-networks. Of note, different causal methods require different sample sizes to achieve similar effects. For example, DBS is the most focal causal method which reduces the number of participants needed to map symptom-networks compared to TMS and lesions. DBS and TMS studies also generally include pre and post intervention symptom data which more tightly links symptoms to circuit modulation, reducing the number of participants needed compared to lesion studies. In addition to large sample sizes, the use of high spatial resolution localization methods also improves precision. With lesions, high spatial resolution can be achieved with careful manual tracing of the original CT/MRI scan, while precision is reduced if the lesion was traced based on a 2-dimensional picture from a case report. With TMS sites, spatial resolution can be maximized with MRI neuronavigation (53), while precision is reduced if non-neuroimaging methods (e.g., scalp measurements) are used. With DBS sites, spatial resolution can be maximized with postoperative lead localization within the targeted nucleus (54,55), and precision is reduced if different nuclei are compared with one another (56,57). We downgraded symptom-networks for imprecision if they were derived from small samples or used low spatial resolution methods to localize lesion/brain stimulation target sites.

Publication bias refers to the overrepresentation of positive results in the published literature. Pre-registration counters publication bias by restricting analytical freedom, reducing the probability of false positive results. To our knowledge, no causal mapping studies have pre-registered their analytical plans to date, and a formal analysis of publication bias in this literature has not been performed. In lieu of clear evidence pertaining to publication bias, we did not evaluate symptom-networks using this criterion. However, we emphasize the lack of pre-registration as a general limitation of causal network mapping studies conducted to date and encourage formal assessment of publication bias in this growing literature.

A large effect magnitude is an evidence strengthening factor. Clinical effects are particularly relevant for evaluating the translational value of a symptom-network. Clinical effects can be tested retrospectively in a number of ways, including by assessing the degree to which TMS/DBS site connectivity to a symptom network predicts symptom change. They can also be assessed prospectively by applying stimulation to a symptom-network target in a randomized trial.

Sometimes an effect may be underestimated by study design features that suppress its observable magnitude. The presence of a significant result despite such suppression increases confidence in an observed effect and is therefore another evidence strengthening factor. Like other synthesis methods, causal network mapping commonly involves aggregating studies that differ in key ways, including the patient populations sampled and the way their symptoms are measured. Although these differences plausibly induce noise and decrease the observable magnitude of an effect, they are not typically considered sufficient to meet the threshold for effect suppression. However, aggregating multiple types of causal evidence (e.g., lesions and TMS sites) into one symptom-network has been shown to reduce effect sizes, whereby the effect size of a depression network generated by one type of causal data was larger (10) than the effect size of a depression networks generated by multiple types (12). Therefore, we upgraded symptom-networks generated by more than one type of causal data.

A final evidence strengthening factor is a dose-response gradient, whereby dose strength predicts clinical effects. A brain stimulation target site with stronger connectivity to a given symptom-network would putatively deliver a stronger stimulation dose to that network. If brain stimulation site connectivity to the network predicts symptom change that would therefore support a dose-response gradient. Another way to demonstrate a dose-effect is to show that the electrical-field induced by an established TMS protocol overlaps with areas showing strong connectivity to a given network. Dose-response gradients can also be assessed via prospective TMS/DBS RCTs that apply stimulation to optimized targets while varying stimulation parameters (i.e., number of pulses, sessions, etc.). We upgraded symptom-networks if they demonstrated a dose-response gradient in one of these ways.

The overall strength of evidence supporting a given symptom-network was evaluated by combining information from each GRADE factor into a composite score. Symptom-networks that were not downgraded (or were boosted by an equivalent number of levels) were rated high quality. Symptom-networks that were downgraded one, two, and three levels were rated moderate, low, and very low quality, respectively.

## Data Availability

All data produced in the present study are available upon reasonable request to the authors

## Supplementary Materials

### I.Symptom-network evaluations

#### Anxiety

<u>Bias:</u> The anxiety network target was not downgraded for bias, as the effects of key confounders were accounted for and the network was shown to be specific to anxiety. Moreover, spatial bias was limited by the inclusion of spatially heterogeneous lesion sites and brain stimulation sites spanning multiple targets. <u>Inconsistency:</u> It was not downgraded for inconsistency due to significant spatial correlations between the lesion, DBS, and TMS networks that comprise it. <u>Indirectness:</u> It was downgraded for indirectness as it was generated from lesion and brain stimulation patients with anxiety symptoms but not necessarily anxiety disorders. However, it should be noted that TMS site connectivity to the anxiety network predicted clinically meaningful changes in anxiety symptoms specifically in depressed patients with anxiety disorders. <u>Imprecision:</u> It was not downgraded for imprecision, as it was derived from several large sample datasets that generally used high resolution techniques to localize lesions/brain stimulation sites. <u>Large effect magnitude:</u> Clinical effect sizes were relatively small, so the network was not upgraded for effect size. <u>Effect suppression</u>: It was upgraded for suppression, as the measured effect size was likely an underestimate due to the integration of multiple causal data-sources with unique confounders. <u>Dose effect:</u> It was upgraded for dose-effect, as greater connectivity to the network predicted greater clinical improvement with therapeutic brain stimulation. <u>Overall grade:</u> The evidence supporting the anxiety network was evaluated as high quality.

#### OCD

<u>Bias:</u> The OCD network target was downgraded for bias because: (1) it was not shown to be specific to OCD versus other disorders/symptoms, and (2) it was derived from DBS sites that were restricted to two small target areas which may have biased results to specific networks. <u>Inconsistency:</u> It was not downgraded for inconsistency due to significant spatial correlations between OCD networks generated from different DBS datasets. <u>Indirectness:</u> The OCD network was generated by brain stimulation data and was therefore not downgraded for indirectness. <u>Imprecision:</u> It was not downgraded for imprecision, as it was derived from a relatively large sample of DBS sites localized using high resolution techniques. <u>Large effect magnitude:</u> Clinical effect sizes were medium, so the network was not upgraded for effect size. <u>Effect suppression</u>: It was not upgraded for suppression, as it was generated using one causal modality. <u>Dose effect:</u> It was upgraded for dose-effect, as greater DBS site connectivity to the network predicted greater clinical improvement. <u>Overall grade:</u> The evidence supporting the OCD network was evaluated as high quality.

#### Addiction

<u>Bias:</u> The addiction network target was not downgraded for bias, as the effects of key confounders were accounted for and the network was shown to be specific to remission of addiction symptoms. Moreover, spatial bias was limited by the inclusion of spatially heterogeneous lesion sites. <u>Inconsistency:</u> It was not downgraded for inconsistency due to significant spatial correlations between addiction networks generated from different lesion datasets. <u>Indirectness:</u> It was downgraded for indirectness because it was generated using lesions and not brain stimulation data. <u>Imprecision:</u> It was not downgraded for imprecision, as it was derived from large sample datasets that generally used high resolution techniques to localize lesions. <u>Large effect magnitude:</u> Clinical effect sizes were not reported, so the network was not upgraded for effect size. <u>Effect suppression</u>: It was not upgraded for suppression, as it was generated using one causal modality. <u>Dose effect:</u> It was upgraded for dose-effect, as the electric-field peaks of multiple TMS coils with established efficacy for addiction were shown to overlap with frontopolar locations with strong connectivity to the addiction network. <u>Overall grade:</u> The evidence supporting the addiction network was evaluated as high quality.

#### PTSD

<u>Bias:</u> The PTSD network target was not downgraded for bias, as the effects of key confounders were accounted for and the network was shown to be specific to protection against PTSD symptoms. Moreover, spatial bias was limited by the inclusion of spatially heterogeneous lesion sites. <u>Inconsistency:</u> It was not downgraded for inconsistency as a significant split-half spatial correlation was demonstrated. <u>Indirectness:</u> It was downgraded for indirectness because it was generated using lesions and not brain stimulation data. <u>Imprecision:</u> It was not downgraded for imprecision, as it was derived from a large sample dataset that used high resolution techniques to localize lesion sites. <u>Large effect magnitude:</u> Clinical effect sizes were not reported, so the network was not upgraded for effect size. <u>Effect suppression</u>: It was not upgraded for suppression, as it was generated using one causal modality. <u>Dose effect:</u> It was upgraded for dose-effect, as greater TMS induced connectivity changes within the network predicted greater PTSD symptom reduction. <u>Overall grade:</u> The evidence supporting the PTSD network was evaluated as high quality.

#### Pain

<u>Bias:</u> The pain network target was not downgraded for bias, as the effects of key confounders were accounted for and the network was shown to be specific to pain. Moreover, spatial bias was limited by the inclusion of spatially heterogeneous lesion sites. <u>Inconsistency:</u> It was not downgraded for inconsistency due to significant spatial correlations with a network generated from an independent sample of pain causing lesions. <u>Indirectness:</u> It was downgraded for indirectness because it was generated using lesions and not brain stimulation data. <u>Imprecision:</u> It was downgraded for imprecision, as it was derived from a relatively small number of pain causing lesions localized using 2d case report images. However, it should be noted that pain causing lesions from the cross-validation dataset were localized using original CT scans. <u>Large</u> <u>effect magnitude:</u> Clinical effect sizes were not reported, so the network was not upgraded for effect size. <u>Effect suppression</u>: It was not upgraded for suppression, as it was generated using one causal modality. <u>Dose effect:</u> The network was upgraded for dose effect, as TMS targets that intersected the network were shown to yield stronger effects on pain and individualized TMS electric field strength within the pain network was shown to predict pain response. <u>Overall grade:</u> The evidence supporting the pain network was evaluated as moderate quality.

#### Psychosis

<u>Bias:</u> The psychosis network target was not downgraded for bias, as the effects of key confounders were accounted for and the network was shown to be specific to psychosis. Moreover, spatial bias was limited by the inclusion of spatially heterogeneous lesion sites. <u>Inconsistency:</u> It was not downgraded for inconsistency as lesion connectivity to the network predicted psychosis symptoms in an out-of-sample dataset. <u>Indirectness:</u> It was downgraded for indirectness because it was generated using lesions and not brain stimulation data. <u>Imprecision:</u> Although it was generated using 2d case report images, it was not downgraded for imprecision because it was derived from a large sample, and the large dataset used to validate it employed high resolution techniques. <u>Large effect magnitude:</u> Clinical effect sizes were not reported, so the network was not upgraded for effect size. <u>Effect suppression</u>: It was not upgraded for suppression, as it was generated using one causal modality. <u>Dose effect:</u> Dose effect relevant results were not reported, so the network was not upgraded for dose effect. <u>Overall grade:</u> The evidence supporting the psychosis network was evaluated as high quality.

#### Aggression

<u>Bias:</u> The aggression network target was not downgraded for bias, as the effects of key confounders were accounted for and the network was shown to be specific to aggression-related constructs (i.e., criminality). Moreover, spatial bias was limited by the inclusion of spatially heterogeneous lesion sites. <u>Inconsistency:</u> It was not downgraded for inconsistency as lesions causing criminality in an independent dataset were shown to significantly intersect the aggression network. <u>Indirectness:</u> It was downgraded for indirectness because it was generated using lesions and not brain stimulation data. <u>Imprecision:</u> It was not downgraded for imprecision, as it was derived from a large sample dataset using high resolution techniques to localize lesion sites. <u>Large effect magnitude:</u> The reported clinical effects were indirect (i.e., DBS site connectivity to the aggression circuit predicted DBS effects on irritability), so the network was not upgraded for effect size. <u>Effect suppression</u>: It was not upgraded for suppression, as it was generated using one causal modality. <u>Dose effect:</u> It was not upgraded for dose-effect for the same reason that it was not upgraded for large effect magnitude. <u>Overall grade:</u> The evidence supporting the aggression network was evaluated as moderate quality.

#### Emotional Regulation

<u>Bias:</u> The emotional regulation network target was not downgraded for bias, as the effects of key confounders were accounted for and the ventral portion of the network was specifically linked to emotional regulation. Moreover, spatial bias was limited by the inclusion of spatially heterogeneous lesion sites. <u>Inconsistency:</u> It was not downgraded for inconsistency as an independent sample of lesions causing emotional regulation related dysfunction (i.e., mania, criminality, depression) were shown to significantly intersect it. <u>Indirectness:</u> It was downgraded for indirectness because it was generated using lesions and not brain stimulation data. <u>Imprecision:</u> It was not downgraded for imprecision, as it was derived from a large dataset using high resolution techniques to localize lesion sites. <u>Large effect magnitude:</u> Clinical effect sizes were not reported, so the network was not upgraded for effect size. <u>Effect suppression</u>: It was not upgraded for suppression, as it was generated using one causal modality. <u>Dose effect:</u> Dose effect relevant results were not reported, so the network was not upgraded for dose effect. <u>Overall grade:</u> The evidence supporting the emotional regulation network was evaluated as moderate quality.

#### Mania

<u>Bias:</u> The mania network target was not downgraded for bias, as the effects of key confounders were accounted for and the network was shown to be specific to mania. Moreover, spatial bias was limited by the inclusion of spatially heterogeneous lesion sites. <u>Inconsistency:</u> It was not downgraded for inconsistency because mania causing lesions from an independent dataset overlapped with the network significantly more than control lesions. <u>Indirectness:</u> It was downgraded for indirectness because it was generated using lesions and not brain stimulation data. <u>Imprecision:</u> It was downgraded for imprecision, as the majority of the lesions used to derive and validate (41/56) it were localized using 2d case report images. Moreover, the network was generated/validated from a relatively small number of mania causing lesions. <u>Large effect</u> <u>magnitude:</u> Clinical effect sizes were not reported, so the network was not upgraded for effect size. <u>Effect suppression</u>: It was not upgraded for suppression, as it was generated using one causal modality. <u>Dose effect:</u> TMS and DBS data provide preliminary evidence for a dose effect. However, these data had noteworthy limitations (i.e., broad, group average targets; small sample) and were therefore deemed not strong enough to justify a dose effect upgrade. <u>Overall grade:</u> The evidence supporting the mania network was evaluated as low quality.

#### Transdiagnostic

<u>Bias:</u> The transdiagnostic network target was downgraded for bias as it was not shown to be specific to psychiatric versus other symptoms. <u>Inconsistency:</u> It was downgraded for inconsistency because it was not subject to within or out-of-sample cross-validation. <u>Indirectness:</u> It was downgraded for indirectness because it was generated using lesions and not brain stimulation data. <u>Imprecision:</u> It was not downgraded for imprecision, as it was derived from a large sample dataset that used high resolution techniques to localize lesion sites. <u>Large</u> <u>effect magnitude:</u> Clinical effect sizes were not reported, so the network was not upgraded for effect size. <u>Effect suppression</u>: It was not upgraded for suppression, as it was generated using one causal modality. <u>Dose effect:</u> Dose effect relevant results were not reported, so the network was not upgraded for dose effect. <u>Overall grade:</u> The evidence supporting the transdiagnostic network was evaluated as very low quality.

#### Criminality

<u>Bias:</u> The criminality network target was not downgraded for bias, as the effects of key confounders were accounted for. Moreover, spatial bias was limited by the inclusion of spatially heterogeneous lesion sites. <u>Inconsistency:</u> It appeared to share a highly similar spatial topography with an out-of-sample lesion criminality network. However, statistically comparisons were not performed. Therefore, it was downgraded for inconsistency. <u>Indirectness:</u> It was downgraded for indirectness because it was generated using lesions and not brain stimulation data. <u>Imprecision:</u> It was downgraded for imprecision, as the lesions used to derive and validate it were localized using 2d case report images. Moreover, it was generated by a small number of lesions (N = 17). <u>Large effect magnitude:</u> Clinical effect sizes were not reported, so the network was not upgraded for effect size. <u>Effect suppression</u>: It was not upgraded for suppression, as it was generated using one causal modality. <u>Dose effect:</u> Dose effect relevant results were not reported, so the network was not upgraded for dose effect. <u>Overall grade:</u> The evidence supporting the criminality network was evaluated as very low quality.

**Supplementary Fig. 1.**
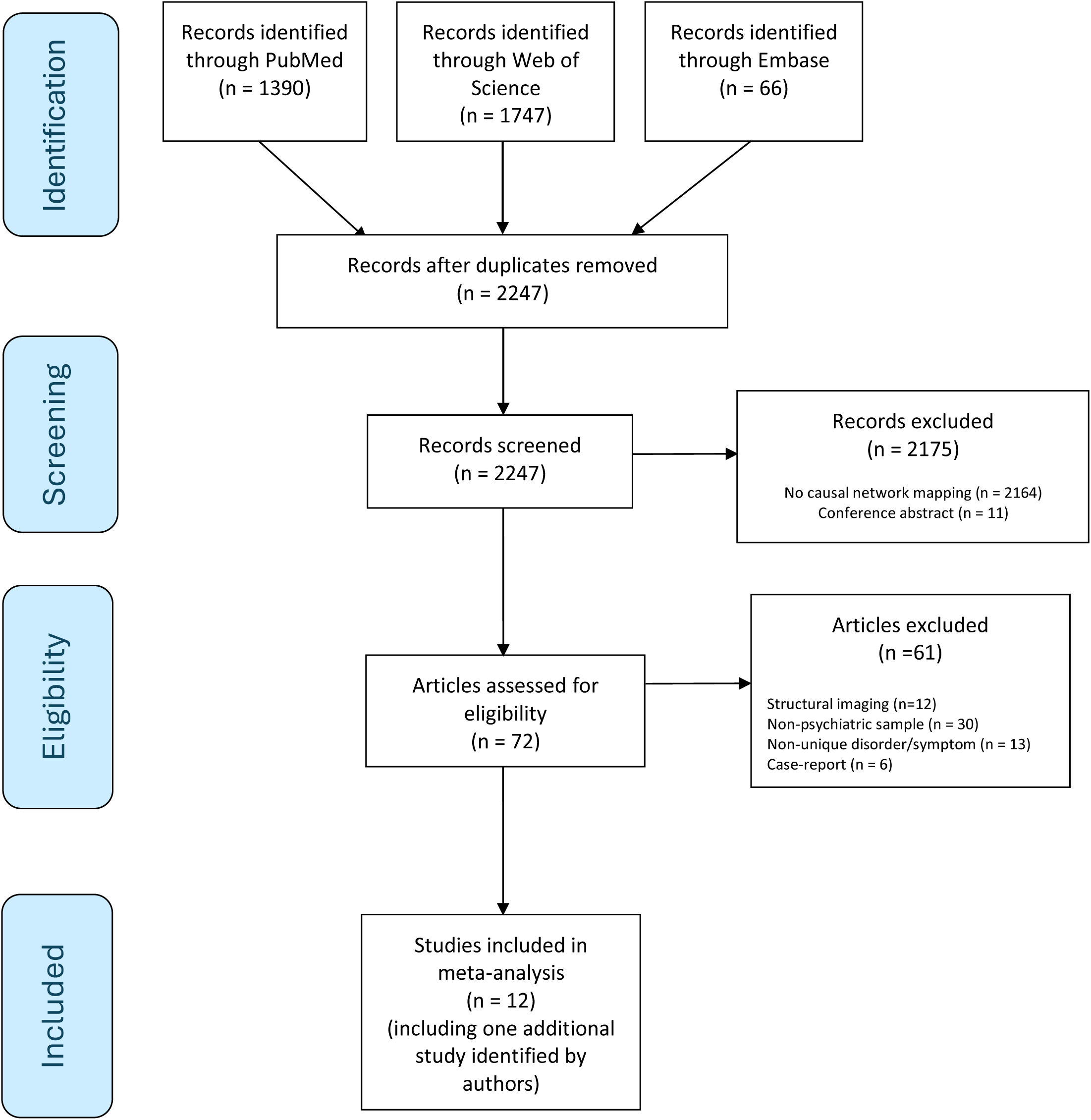
PRISMA Diagram. Flowchart depicting preferred reporting items for systematic reviews and meta-analyses (PRISMA) guided study selection process. For information on PRISMA, see: http://www.prismastatement.org/.

